# Dual-Prep registry: atherectomy Devices and intravascUlAr Lithotripsy for the PREParation of heavily calcified coronary lesions registry, 1-year Results

**DOI:** 10.64898/2025.12.17.25342522

**Authors:** Masato Nakamura, Nehiro Kuriyama, Yutaka Tanaka, Seiji Yamazaki, Tomohiro Kawasaki, Takashi Muramatsu, Kazushige Kadota, Takashi Ashikaga, Akihiko Takahashi, Satoru Otsuji, Kenji Ando, Masaru Ishida, Shigeru Nakamura, Yoshiaki Ito, Raisuke Iijima, Gaku Nakazawa, Junya Shite, Junko Honye, Junya Ako, Hiroyoshi Yokoi, Ken Kozuma, Hiromasa Otake, Kenji Kochi, Tomomi Yamada, Yohei Sotomi

**Author notes:** Address for correspondence: Masato Nakamura, MD, PhD, Division of Minimally Invasive Treatment in Cardiovascular Medicine, Toho University Ohashi Medical Center. Address: 2-22-36, Ohashi, Meguro-ku, Tokyo 153-8515, Japan.

## Abstract

**Background:** Combination therapy with atherectomy and intravascular lithotripsy (IVL) has emerged as a promising strategy for the treatment of severely calcified occlusive coronary lesions, which potentially enhances procedural efficacy without increasing complication risk.

**Methods:** The Dual-Prep Registry is a multicenter, prospective registry designed to evaluate the safety and efficacy of IVL after atherectomy in severely calcified lesions. Combined use was selectively applied when the risk of complications was anticipated to be high with a larger atherectomy burr size, or when it was deemed non-beneficial due to unfavorable guidewire bias. All adverse events were adjudicated by a clinical events committee. Kaplan-Meier analysis was performed to evaluate the primary endpoint of major adverse cardiovascular events (MACE; composite endpoint of cardiac death, myocardial infarction, and target vessel revascularization [TVR]) at 1 year.

**Results:** A total of 118 cases (120 lesions) were enrolled across 20 facilities. Significant comorbidities included diabetes in 56.8% of patients and hemodialysis-dependence in 25.4%. Calcification score after atherectomy was 4.0 in all cases, and calcified nodules were present in 56.4% (core-lab analysis) of cases. One-year follow-up was complete in 99.2% patients. MACE occurred in 7.6% patients at one year (cardiac death 2.5%, myocardial infarction 5.1%, TVR 5.1%) and stent thrombosis was observed in 1 case.

**Conclusions:** Atherectomy followed by IVL resulted in low 1-year rates of MACE, TVR, and stent thrombosis in patients with severely calcified coronary lesions. This approach may be considered for lesions where an “IVL-first” strategy is difficult to apply.

Japan Registry of Clinical Trials: jRCT1032230384. URL: https://jrct.mhlw.go.jp

**A Clinical Perspective:**

1) What Is New?

- Elective combined use of IVL and atherectomy resulted in low 1 year MACE and TLR.
- The incidence of MACE was higher in cases with greater residual stenosis after the procedure and a larger baseline reference vessel diameter.
2) What Are the Clinical Implications?

- Combining IVL with atherectomy may serve as an effective treatment strategy in cases where IVL-first approaches are difficult to apply.
- Severe calcified lesions that are presumed to be unresponsive to RA/OA treatment or carry a high risk of RA/OA complications may be good candidates for this strategy.

## Introduction

Calcified coronary lesions remain a major barrier to percutaneous coronary intervention (PCI), determining not only short-term but also long-term outcomes.^1, 2^ Since these outcomes cannot be overcome solely by advances in final treatment devices such as drug-eluting stents (DES) and drug-coated balloons (DCB), the importance of pre-DES/DCB lesion preparation as a preliminary step has been emphasized, and lead to the exploration of a range of approaches. One of these is intravascular lithotripsy (IVL), a recently developed lesion preparation device for calcified lesions. It uses ultrasonic acoustic pressure waves to fragment calcification in highly calcified lesions, allowing subsequent optimal vessel expansion in both coronary and peripheral vascular beds. Previous findings indicate that this technology offers advantages over atherectomy, including lower risk of vascular injury and distal embolism from microparticles, and potentially better stent expansion compared to other lesion preparation strategies.^3, 4, 5^ Its use in clinical practice has reportedly increased over time, suggesting high device safety.^6, 7^ Furthermore, registry data indicate a concurrent trend of increasing combined use with atherectomy over time, likely reflecting the expectation that combined therapy serves as a complementary and synergistic treatment strategy which enhances efficacy without increasing risk. This is because while atherectomy carries a risk of distal embolization and is ineffective in cases with unfavorable guidewire bias, IVL sometimes has the disadvantage of low device crossability. The Dual Prep Registry evaluated the concept that combining atherectomy with IVL offers benefits, and demonstrated that the addition of IVL after atherectomy was effective in cases where the sizing of atherectomy burr size was deemed likely ineffective, or where there were concerns about complication risks.^8^ However, reports on the long-term efficacy of IVL after atherectomy remain scarce and the question remains unanswered.

Here, we report the one-year clinical outcomes of DUAL-PREP, a trial based on the Dual-Prep Registry.

### Study design

DUAL-PREP is a prospective, multicenter, single-arm study designed to assess the safety and efficacy of combination use of atherectomy devices and IVL before DES deployment in patients with severely calcified lesions. Severity of calcification was initially evaluated angiographically. A calcification score of 3 or greater prior to IVL was a mandatory requirement, and was subsequently confirmed by intravascular imaging. The details of the study protocol have been published previously.^8^ In brief, enrollment required: 1) age 18 years or older; 2) consent to participate in the study; and 3) the presence of severely calcified lesions for which treatment with IVL after other atherectomy devices may be preferable, based on imaging findings. Exclusion criteria included: 1) participation in other clinical trials that may affect the results; and 2) ineligibility for treatment with atherectomy or IVL. Atherectomy devices eligible for use included the ROTAPRO^TM^ Rotational Atherectomy System (Boston Scientific, Marlborough, Massachusetts) and Diamondback 360^TM^ Coronary Orbital Atherectomy System (Abbott Vascular, Santa Clara, California), while the Shockwave C^2^ Coronary IVL System (Shockwave Medical, Santa Clara, California) was utilized for IVL. The study protocol was approved by central review (Institutional Review Board of Toho University Ohashi Hospital Ethics Board H23024 H23 May 22, 2023). A flow chart of the present study is shown in Figure 1. RA/OA was performed at the operator’s discretion in a standard fashion.^9^ After atherectomy (RA/OA), IVL was used when a calcified lesion was considered inadequately pretreated but further atherectomy was inappropriate, e.g. slow flow, deep calcification, or when guidewire bias limited atherectomy effectiveness. Finally, DES was deployed after IVL and any subsequent postdilatation. Imaging evaluation of coronary lesions with OCT/OFDI was scheduled at least four times: before lesion instrumentation, after RA/OA, after IVL, and after stent deployment. Intravascular imaging was used to assess not only the severity of calcified lesions but also the appropriateness of device selection, such as guidewire bias and stent expansion etc.

**Figure 1.**
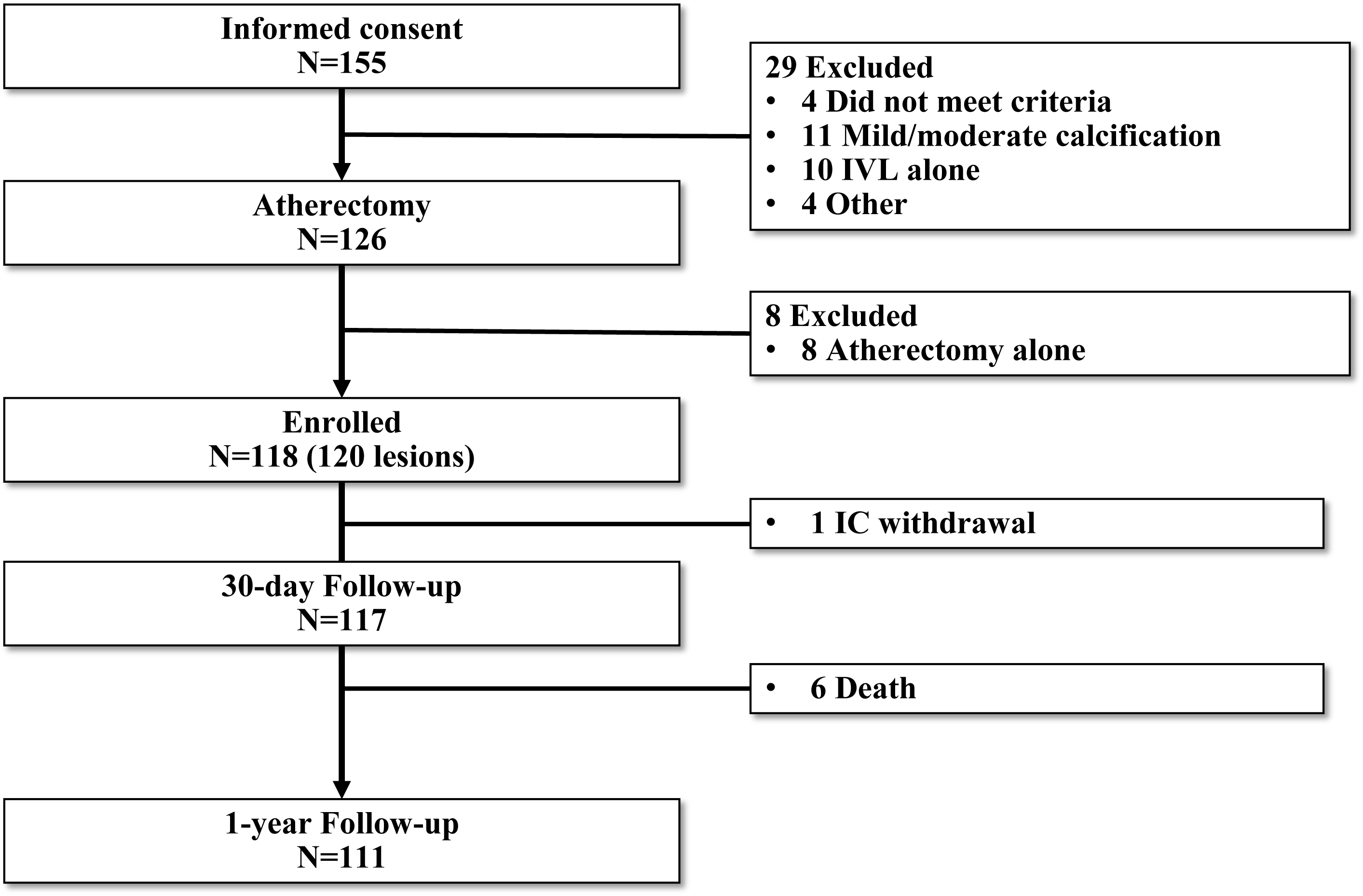
Study flow chart. Patients were enrolled from 20 sites in Japan through November 2023 to June 2024. 1 year follow-up rate was 99.2%. IVL: intravascular lithotripsy

### Study endpoints

The primary endpoint of the study was major adverse cardiac events (MACE) at 1 year after the procedure, defined as a composite of cardiovascular death, myocardial infarction (MI) or clinically driven target vessel revascularization (CD-TVR). MI was classified into periprocedural MI and spontaneous MI, with the former adjudicated according to the Society for Cardiovascular Angiography and Interventions (SCAI) definition^10^ and the latter by the 4th Universal definition.^11^ TLF, defined as a composite of cardiac death, MI, and target lesion revascularization (TLR), was also evaluated. Procedural success was defined as residual stenosis <50% after stenting without MACE during hospitalization. Angiographic success was defined as successful IVL crossing, balloon expansion & therapy delivery, residual stenosis ≤30% after stenting, and no serious angiographic complications (core laboratory assessment). Angiograms and intravascular imaging (OCT/OFDI) were independently adjudicated by a core lab (Micron, Inc, Osaka, Japan), and clinical events were independently adjudicated by a Clinical Events Committee (CEC).

### Statistical Analysis

Continuous variables were expressed as mean±SD. Categorical variables were summarized as frequencies and proportions. Kaplan-Meier analysis was performed to estimate cumulative event-free rate of MACE, TLF, and component of MACE at 1-year follow-up. 95% confidence interval (CI) was calculated by Greenwood’s formula. Furthermore, Brookmeyer and Crowley’s method was used to calculate the CI for median survival time. Factors associated with MACE were evaluated using univariate logistic regression analysis with Firth’s penalized likelihood method. All tests were two-sided, and p values <0.05 were considered statistically significant. Analyses were performed by an independent biostatistician using SAS version 9.4 statistical analysis software (SAS Institute Inc., Cary, NC, USA).

## Results

### Subjects and procedure

Informed consent was obtained from 156 patients at 20 participating centers between November 2023 and June 2024. From this cohort, 28 cases that did not undergo atherectomy and 8 cases that underwent atherectomy alone without concomitant IVL were not registered in this registry. The primary reason for exclusion was insufficient degree of calcification severity on intravascular imaging rather than failure to meet the registration criteria. Consequently, 118 cases (120 lesions) were analyzed in this registry. All patients were enrolled at the time of attempted treatment with the IVL system after atherectomy, regardless of whether the IVL catheter reached the lesion or not. A flow chart of the present study is shown in Figure 1.

Patient demographics and lesion characteristics are depicted in Table 1. Mean age was 75.8±8.9 years, 70.3% of patients were male, 56.8% had diabetes mellitus and 25.4% were on hemodialysis. 91.5% had de novo coronary artery lesions and were primarily patients with chronic coronary syndrome (89.8%). The principal treated vessel was the left anterior descending artery (64.2%) and mean reference vessel diameter was 2.67±0.69 mm, with mean lesion length 34.3±15.2 mm. Procedural factors are shown in Table 2. A transradial approach was applied in 63.5% of cases. RA was performed in 83.9% of patients with a mean burr size of 1.57±0.20 mm. The remaining 16.9% underwent OA. The calcification score of lesions after atherectomy remained at 4.0 in all cases. Additional IVL after atherectomy was performed in 42.4% of cases due to safety concerns; in 60.2% of cases because additional atherectomy was not expected to be effective; and in 1.7% of cases for other reasons (multiple responses allowed). In all patients, the IVL was successfully traversed to the target lesion site. The IVLs were primarily 2.5 mm and 3.5 mm in diameter, and used in 36.7% and 44.2% of cases, respectively. A DES was implanted in all cases, with a mean diameter of 3.19±0.51 mm and length of 36.3±16.0 mm. DES postdilatation was performed in 79.2% of cases, with a mean maximum balloon diameter of 3.45±0.58mm mm and mean inflation pressure of 17.9±4.6 atmospheres. Procedural success was achieved in 117/120 lesions (97.5%; 92.9-99.5). Following PCI, 68 patients (57.6%) continued dual antiplatelet therapy for one year, while 24.6% were prescribed P2Y12 inhibitor monotherapy (Figure S1).

**Table 1:**
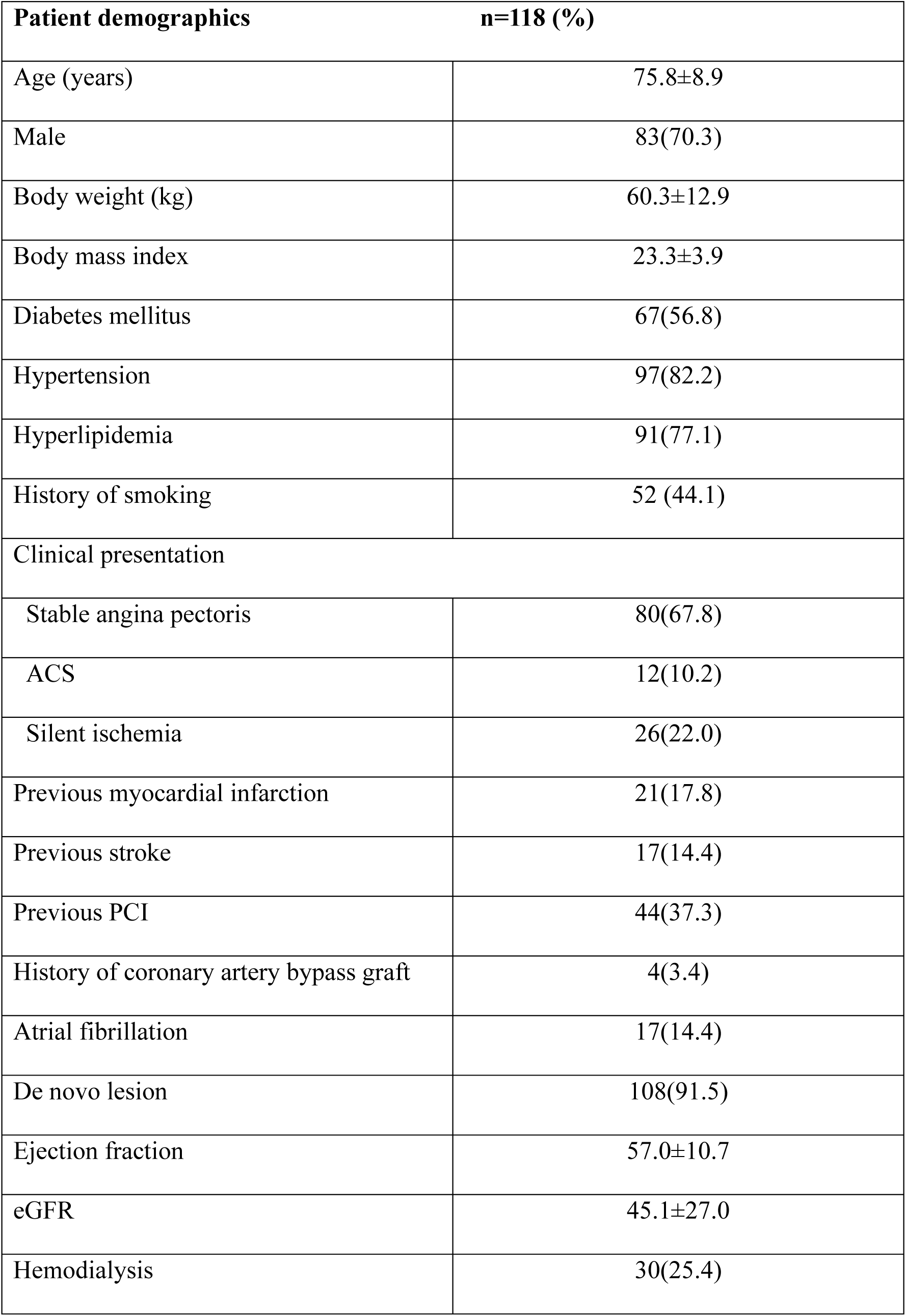
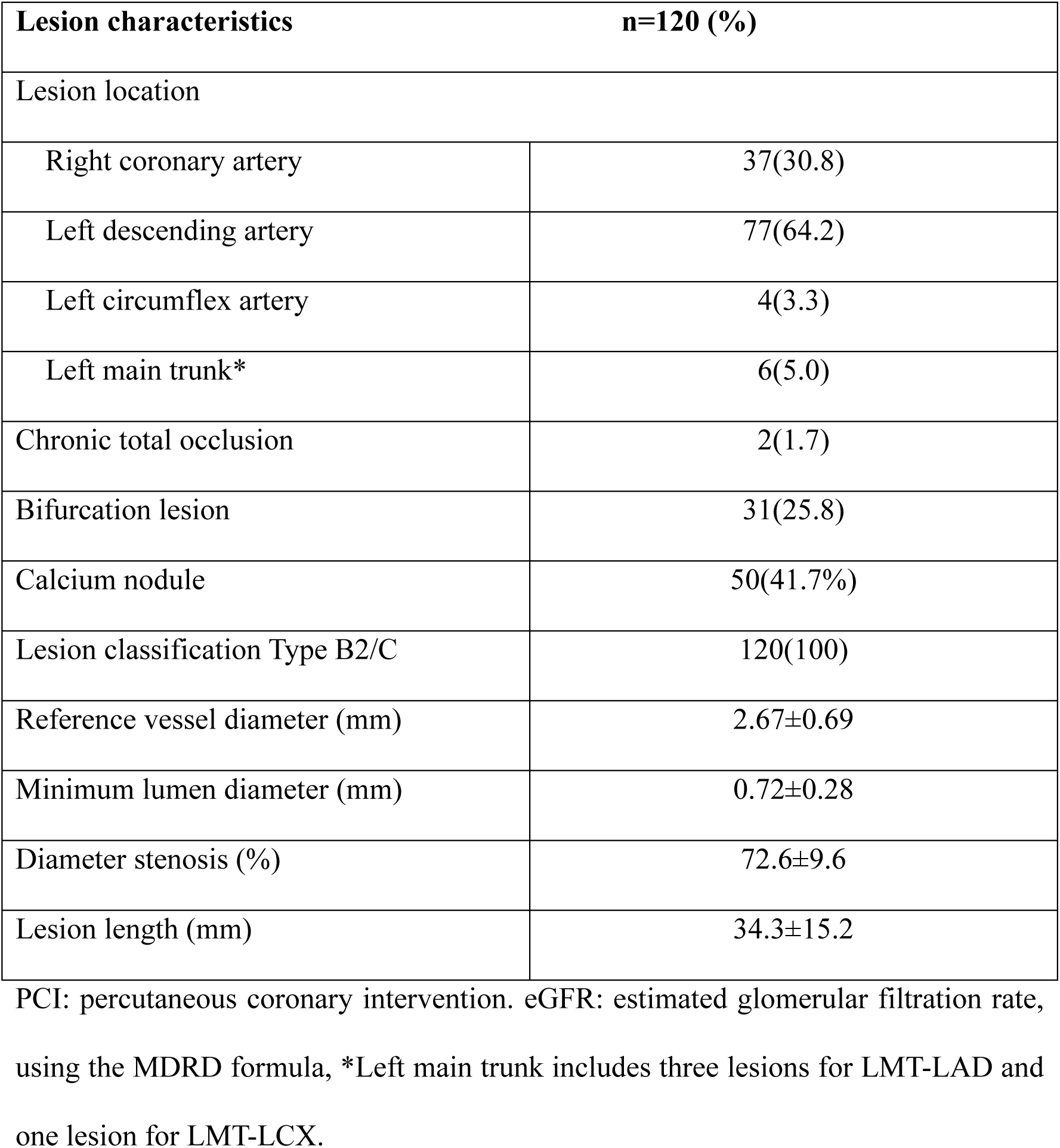
Patient demographics and lesion characteristics.

| **Patient demographics** |  | **n=118 (%)** |
| --- | --- | --- |
| Age (years) |  | 75.8±8.9 |
| Male |  | 83(70.3) |
| Body weight (kg) |  | 60.3±12.9 |
| Body mass index |  | 23.3±3.9 |
| Diabetes mellitus |  | 67(56.8) |
| Hypertension |  | 97(82.2) |
| Hyperlipidemia |  | 91(77.1) |
| History of smoking |  | 52 (44.1) |
| Clinical presentation |  |  |
| Stable angina pectoris |  | 80(67.8) |
| ACS |  | 12(10.2) |
| Silent ischemia |  | 26(22.0) |
| Previous myocardial infarction |  | 21(17.8) |
| Previous stroke |  | 17(14.4) |
| Previous PCI |  | 44(37.3) |
| History of coronary artery bypass graft |  | 4(3.4) |
| Atrial fibrillation |  | 17(14.4) |
| De novo lesion |  | 108(91.5) |
| Ejection fraction |  | 57.0±10.7 |
| eGFR |  | 45.1±27.0 |
| Hemodialysis |  | 30(25.4) |

| Lesion characteristics |  | n=120 (%) |
| --- | --- | --- |
| Lesion location |  |  |
| Right coronary artery |  | 37(30.8) |
| Left descending artery |  | 77(64.2) |
| Left circumflex artery |  | 4(3.3) |
| Left main trunk* |  | 6(5.0) |
| Chronic total occlusion |  | 2(1.7) |
| Bifurcation lesion |  | 31(25.8) |
| Calcium nodule |  | 50(41.7%) |
| Lesion classification Type B2/C |  | 120(100) |
| Reference vessel diameter (mm) |  | 2.67±0.69 |
| Minimum lumen diameter (mm) |  | 0.72±0.28 |
| Diameter stenosis (%) |  | 72.6±9.6 |
| Lesion length (mm) |  | 34.3±15.2 |

PCI: percutaneous coronary intervention. eGFR: estimated glomerular filtration rate, using the MDRD formula, \*Left main trunk includes three lesions for LMT-LAD and one lesion for LMT-LCX.

**Table 2:** Procedural characteristics.

| **Characteristic** |  |
| --- | --- |
| Radial approach | 75(63.6) |
| Transfemoral approach | 38(32.2) |
| Brachial approach | 5(4.2) |
| Rotational atherectomy* | 99(83.9) |
| Step up of burr size | 10(8.5) |
| Used burr size |  |
| 1.25 mm | 17(17.2) |
| 1.5 mm | 60(60.6) |
| 1.75 mm | 23(23.2) |
| 2.0 mm | 9(9.1) |
| Orbital atherectomy | 20(16.9) |
| IVL treatment |  |
| Number of catheters per case | 1.10±0.30 |
| 2.5 mm | 44(36.7) |
| 3.0 mm | 53(44.2) |
| 3.5 mm | 23(19.2) |
| 4.0 mm | 12(10.0) |
| Total number of pulses | 76.5±22.9 |
| Side branch protection | 29(24.2) |
| Postdilatation before stent deployment | 51(42.5) |
| Modified balloon | 32(62.7) |
| Non-compliant balloon | 20(39.2) |
| Max balloon size (mm) | 2.96±0.48 |
| Max dilatation pressure (atm) | 16.5±4.8 |
| Drug eluting stent |  |
| Stent diameter (mm) | 3.19±0.51 |
| Total stent length (mm) | 36.3±16.0 |
| Post-stent dilatation | 95(79.2) |
| Max balloon size (mm) | 3.45±0.58 |
| Max dilatation pressure (atm) | 17.9±4.6 |
| OCT/OFDI use as a guidance of PCI | 109 (90.8) |
| IVUS use as a guidance of PCI** | 19 (16.5) |
| Procedural success | 117 (97.5%) |

IVL: intravascular lithotripsy, IVUS; intravascular ultrasound, OCT/OFDI: optical coherence tomography/optical frequency domain imaging
\*1 case was treated with RA followed by OA.
\*\*Both IVUS and OCT/OFDI were applied to guide PCI in 8 lesions.

### 1-year outcomes

The 1-year follow-up rate was 99.2%. The Kaplan-Meier estimates for the 1-year MACE-free and TLF-free rates were both 92.3% (95%CI: 85.8–95.9%), and the 1-year TLR-free rate was estimated to be 94.8% (95%CI: 88.9–97.7%) (Figure 2). The incidence of MACE components and secondary endpoints is shown in Table 3 and Figure S2, respectively. All-cause death occurred in 5.1%, of which 2.5% were cardiac deaths. All TVRs (5.1%) were related to the target lesion. MI occurred in 5.1% of patients, including 1.7% procedure-related MIs. Stent thrombosis occurred in 1 case.

**Figure 2.**
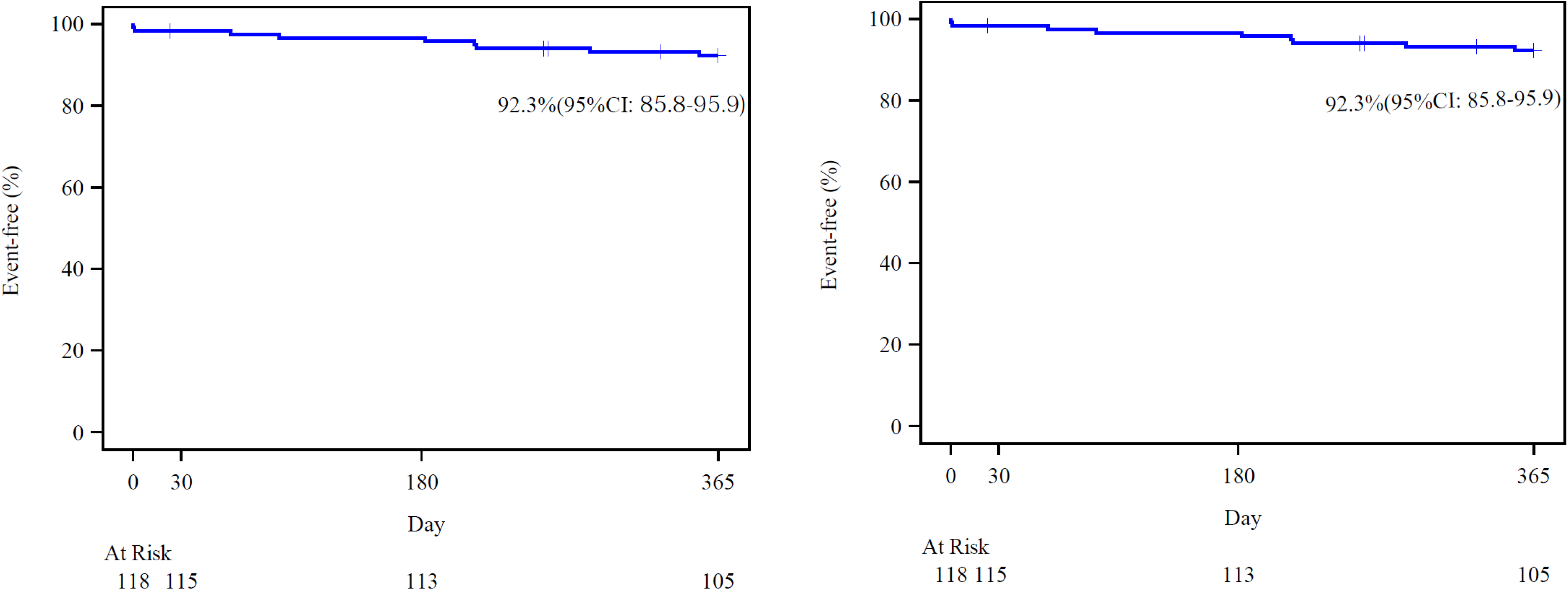
The Kaplan-Meier curve for 1-year MACE-free and TLF-free rates.

**Table 3:** 1-year outcomes.

|  | n (%) |
| --- | --- |
| MACE 1 year | 9 (7.6) |
| Cardiac death | 3 (2.5) |
| MI | 6 (5.1) |
| CD-TVR | 6 (5.1) |
| TLF | 9 (7.6) |
| Stent thrombosis | 1 (0.8) |
| Definite | 1 (0.8) |
| Probable | 0 (0) |
| All death | 6 (5.1) |
| Cardiovascular death | 4 (3.4) |
| Non-CV death | 2(1.7) |
| All MI | 6 (5.1) |
| Periprocedural MI | 2 (1.7) |
| Spontaneous MI | 4 (3.4) |
| All revascularization | 9 (7.6) |
| CD-TLR | 6 (5.1) |
| Non-target-vessel-related TVR | 3 (2.5) |

MACE: major adverse cardiac event, MI: myocardial infarction, TVR: target vessel revascularization.
TLR: target lesion revascularization.

### Factors associating with MACE

Table 4 shows a univariate logistic regression analysis for MACE. Actual incidence rates in various subgroups are shown in Table S1. Both % residual stenosis after stent placement and increased reference vessel size showed a significant positive association with MACE, while other factors such as procedural characteristics and patient background were not associated with MACE. A 5% increase in residual stenosis and 1-mm increase in reference vessel diameter were each associated with a 2.3-fold increase in MACE.

**Table 4:** Univariate logistic regression analysis with Firth’s penalized likelihood approach.

|  | OR | 95% CI | p-Value |
| --- | --- | --- | --- |
| Age (each 5 year increase) | 1.53 | 0.96-2.42 | 0.072 |
| Male | 0.31 | 0.08-1.18 | 0.087 |
| Body weight (each 5 kg increase) | 0.91 | 0.70-1.19 | 0.503 |
| DM | 2.46 | 0.55-10.93 | 0.239 |
| HL | 0.33 | 0.09-1.27 | 0.108 |
| Smoking | 0.80 | 0.22-2.98 | 0.744 |
| Dialysis | 2.58 | 0.68-9.80 | 0.165 |
| eGFR (each 10 ml/min/1.73m<sup>2</sup> increase) | 0.82 | 0.64-1.05 | 0.118 |
| LAD | 0.44 | 0.12-1.65 | 0.223 |
| Ostial | 1.68 | 0.38-7.53 | 0.497 |
| Reference (each 1 mm increase) | 2.27 | 1.05-4.90 | 0.037 |
| Pre MLD (each 1 mm increase) | 4.92 | 0.63-38.6 | 0.130 |
| Pre % DS (each 5% increase) | 1.06 | 0.75-1.49 | 0.753 |
| Post MLD (each 1 mm increase) | 1.60 | 0.52-4.89 | 0.413 |
| Post % DS (each 5% increase) | 2.34 | 1.28-4.30 | 0.006 |
| Burr to Artery ratio (each 0.1 increase) | 0.64 | 0.35-1.17 | 0.150 |
| IVL to vessel ratio (each 0.1 increase) | 0.91 | 0.68-1.22 | 0.522 |
| Stent to vessel ratio (each 0.1 increase) | 0.84 | 0.61-1.15 | 0.269 |
| Eruptive calcified nodule | 1.78 | 0.43-7.33 | 0.422 |
| Non-eruptive calcified nodule | 0.50 | 0.07-3.43 | 0.478 |
| Min stent expansion index (each 10 increase) | 1.01 | 0.64-1.57 | 0.977 |
| Asymmetry index (each 0.1 increase) | 1.08 | 0.72-1.62 | 0.708 |
| Min eccentricity index (each 0.1 increase) | 0.78 | 0.40-1.53 | 0.476 |
| Min stent area (each 1 mm<sup>2</sup> increase) | 1.19 | 0.92-1.55 | 0.183 |

## Discussion

This is the first prospective study to evaluate 1-year outcomes of follow-on use of IVL after atherectomy for severely calcified coronary lesions under intravascular imaging guidance. Results showed that the estimated freedom from MACE at 1 year was 92.3% (95%CI: 85.8–95.9%) and the estimated 1-year TLR-free rate was 94.8% (95%CI: 88.9–97.7%). Further, factors associated with MACE were residual % stenosis and reference vessel diameter. These findings suggest that the combination of IVL with atherectomy may be an effective treatment strategy in cases where IVL-first approaches are difficult to apply.

The conceptual efficacy of IVL has been demonstrated in the pivotal DISRUPT trial series.^12, 13, 14, 15^ In daily clinical practice, however, IVL is used for a broader range of indications. A recent report ^12^ on trends in atherectomy and IVL use among PCI patients in the US noted that IVL usage increased from 0.04% to 2.57% between January 2021 and June 2022.^6^ Among these cases, only 15.6% met the inclusion or exclusion criteria of the pivotal trials. Cases not meeting the approval trial criteria were primarily acute coronary syndrome (ACS) cases, in-stent restenosis, and combined treatments with atherectomy. This may reflect expectations for IVL and confidence in its safety, while also suggesting that cases in which IVL or atherectomy alone is difficult to perform remain a challenge. Furthermore, reports from the US CathPCI Registry also indicate that combined therapy has increased over time alongside the rise in IVL use.^7^ Although outcomes are primarily evaluated in small, retrospective studies assessing short-term results, a meta-analysis of 12 observational studies involving 720 patients reported a procedural success rate of 93%.^16^ The authors concluded that the combination of atherectomy and IVL for calcified coronary lesions is feasible with a high procedural success rate. However, this report also included numerous IVL uses in different situations, such as upfront use or bail-out scenarios, requiring caution in interpretation.

The DUAL-Prep Registry addresses these limitations. This prospective registry study evaluated the additive effects of combining RA and IVL in cases where increased burr size after initial RA/OA therapy was considered unlikely to be effective or where a high risk of complications was anticipated. Additionally, intravascular imaging was used to assess calcification severity, confirming that all cases exhibited severe calcification even after RA/OA and required additional interventional treatment for calcified lesions. Thus, this study represents an outcome assessment of combined atherectomy and IVL therapy based on a pragmatic concept. A previous acute study reported a 30-day MACE avoidance rate of 98.3% (CI: 94.0-99.8%),^8^ which is favorable compared to outcomes from treatment strategies using other devices with a lower complication rate. Thanks to that study, combination therapy with atherectomy and IVL has emerged as a promising strategy for treating severely calcified occlusive coronary lesions, which potentially enhances procedural efficacy without increasing complication risk. Although the Dual-Prep Registry subsequently demonstrated high procedural initial success with IVL following atherectomy, long-term clinical outcomes remained uncertain. The present study sought to address this question.

In this longer-term follow-up, estimated freedom from MACE at 1 year was favorable, at 92.3%. In a study on long-term outcomes reported for 114 cases of combination therapy conducted in Europe from May 2019 to December 2023,^17^ the reported one-year MACE of 9% appears consistent with the current results; however, it should be noted that the prior study was retrospective and had a longer registration period (5 years) compared to this trial (8 months). Furthermore, the elective combined approach rate for RA/OA-IVL was 100% in this study versus 57% in the retrospective report, and the one-year follow-up rate was 97% versus 67%. Therefore, the Dual-Prep registry is the first study to demonstrate the 1-year efficacy of combination therapy without these potentially confounding factors, and in a prospective fashion. In particular, 42% of our enrolled cases were deemed unsuitable for IVL because the imaging catheter itself could not be passed through, 25.4% were dialysis patients, and 56.4% had calcified nodules (CN) requiring RA/OA. We therefore consider that the MACE incidence of 7.6% and TLR incidence of 5.1% in this study to be favorable outcomes.

Moreover, the MACE outcomes in this study occurred at a lower rate than the long-term MACE rate associated with RA therapy alone (17.7% at 15-month follow-up) or the 9-month MACE rate of 24.2% in the ROTAXUS trial.^18, 19^ The most likely reasons for the superiority of MACE rate compared to RA alone likely include the strategic use of smaller burr-vessel ratios paired with the use of IVL to complete the procedure. This may contribute to a reduction in in-hospital events, including procedural complications. Furthermore, the synergistic effects of this combination strategy also likely contribute. In this regard, on comparison of results with the stent expansion index using OCT, Dual-Prep demonstrated a better stent expansion index than the other treatment strategies. An inadequate stent expansion index suggests suboptimal stent expansion and is known to be an important determinant of long-term stent failure. Therefore, it is reasonable to consider that the favorable OCT analysis results of DUAL-Prep directly contributed to improved long-term outcomes.

### Factors associated with MACE

Factors associated with MACE included baseline reference vessel diameter and postprocedural % stenosis. The incidence of MACE was significantly higher as both factors increased. This finding is partially consistent with the BENELUX-IVL registry, which reported clinical and technical predictors of MACE after coronary IVL.^20^ In the BENELUX-IVL registry, multivariate analysis revealed that procedural complications, chronic total occlusion, in-stent restenosis, plaque modification after IVL, and greater post procedural residual stenosis were associated with MACE. Compared to their study, the present study used pre-IVL plaque modification in all cases, and excluded ISR cases. Furthermore, the in-hospital complication rate in this study was lower than that in the BENELUX-IVL Registry, with the proportion of ACS of 10.2% and 42%, respectively. These studies can be considered to have produced essentially similar results given the baseline differences in the populations studied. These findings reaffirm residual stenosis as a universal indicator and emphasize its importance as a procedural guide tool. Another factor is large vessels: luminal gain is often greater in larger vessels than small vessels, potentially leading physicians to consider the procedure adequate even when residual stenosis is suboptimal/inadequate. As vessel diameter increases, the thickness of calcification may also increase, resulting in more inadequate modification and consequently larger residual stenosis in large vessels. Notably, CN were not a determinant of outcome in this study. Similarly, Ali et al. reported a pooled analysis using OCT data from the Disrupt CAD trials,^13^ in which CN were identified in 18.7% (29/155) of lesions among 155 subjects, and IVL use resulted in equivalent stent expansion and lumen enlargement in both CN and non-CN lesions. The Dual-Prep Registry OCT analysis yielded similar results, showing no difference in stent expansion based on CN presence. Adequate stent expansion is a fundamental requirement for favorable long-term outcomes, and is therefore likely reflected in the long-term freedom from MACE observed. Although no significant association was observed, the directionality of the association with MACE differed between eruptive CN and non-eruptive CN: eruptive CN showed a stronger association with MACE, consistent with findings suggested by previous reports.^21^

### Limitations

Several important limitations of this trial must be noted. First, this was an open-label, single-arm trial. Comparative studies based on hypotheses are needed to assess the superiority of this strategy over other treatment strategies. Second, the selection of treatment device size and subsequent therapy was left to the treating physician’s judgment and was not based on consistent rules. Third, the frequency of ACS was low, and it is unclear whether efficacy is consistent across all clinical situations. Fourth, while increased healthcare costs are a concern, the balance of cost-effectiveness remains unclear. Fifth, given the risk of overfitting due to the small number of events, multivariate analysis of event-related factors could not be performed, and caution should be exercised in interpreting this analysis. Therefore, these results require validation in future prospective studies. Finally, long-term prognosis over more than 1 year remains unclear.

## Conclusions

Atherectomy followed by IVL resulted in low 1-year rates of MACE, TLR, and stent thrombosis in patients with severely calcified coronary lesions. This approach may be considered for lesions where an “IVL-first” strategy is difficult to apply.

## Data Availability

Data will not be available to others.

## Acknowledgements

The authors thank Yoshito Nakamoto and Kou Tachikawa (Micron, Inc. Osaka, Japan) for their assistance with the core lab analysis, and Asahi Funayama and Kanako Imamura (Micron, Inc. Tokyo, Japan) for study management.

## Conflict of interest statement

The Dual-Prep Registry was sponsored and funded by Shockwave Medical Inc. (Santa Clara, California, USA), which played a role in the study design discussion but was not involved in the collection, management, analysis, or interpretation of the data. Dr. Nakamura Masato has received consulting fees from Shockwave Medical Japan K.K., honoraria from Boston Scientific Japan K.K., Terumo Co. and Shockwave Medical Japan K.K. and endowments from Boston Scientific Japan, K.K. and Terumo Co. Dr. Tanaka has received honoraria from Shockwave Medical Japan K.K. Dr. Kawasaki has received honoraria from Abbott Japan LLC., Boston Scientific Japan K.K., and Terumo Co. Dr. Muramatsu has received honoraria from Abbott Japan LLC., Boston Scientific Japan K.K., Shockwave Medical Japan K.K., and Terumo Co. Dr. Otsuji has received honoraria from Abbott Japan LLC., Boston Scientific Japan K.K., and Shockwave Medical Japan K.K. Dr. Ando has received honoraria from Abbott Japan LLC. and Terumo Co. Dr. Ishida has received honoraria from Abbott Japan LLC., Boston Scientific Japan K.K., and Terumo Co. Dr. Ito has received honoraria from Boston Scientific Japan K.K., Abbott Japan LCC., and Terumo Co. Dr. Iijima has received honoraria from Boston Scientific Japan K.K. and Terumo Co. Dr. Nakazawa has received honoraria from Abbott Japan LLC., Boston Scientific Japan K.K., Terumo Co., and Shockwave Medical Japan K.K. Dr. Ako has received honoraria from Shockwave Medical Japan K.K. Dr. Kozuma has received honoraria from Boston Scientific Japan K.K., and Abbott Japan LLC and is a board member of Cardiovascular Intervention and Therapeutics. Dr. Otake has received grants from Abbott Japan LLC and honoraria from Abbott Japan LCC and Terumo Co. Dr. Sotomi has received grants from Abbott Japan LCC., Boston Scientific Japan K.K., and Terumo Co. Dr. Kuriyama, Dr. Yamazaki, Dr. Kadota, Dr. Ashikaga, Dr, Takahashi, Dr. Nakamura Shigeru, Dr. Shite, Dr. Honye, Dr. Yokoi, Mr. Kochi, and Dr. Yamada have no conflicts of interest to disclose.

## Non-standard Abbreviations and Acronyms

CD-TVR: Clinically driven target vessel revascularization
CEC: Clinical Evaluation Committee
CI: Confidence interval
CN: Calcified nodule
DCB: Drug-coated balloon
DES: Drug eluting stent
IVL: Intravascular lithotripsy
MACE: Major adverse cardiac events
MI: Myocardial infarction
OA: Orbital atherectomy
OCT: Optical coherence tomography
OFDI: Optical frequency domain imaging
PCI: Percutaneous coronary intervention
RA: Rotational atherectomy
SCAI: Society for Cardiovascular Angiography and Interventions
TLF: Target lesion failure
TLR: Target lesion revascularization
TVR: Target vessel revascularization

## Notes

### Clinical Trial

Japan Registry of Clinical Trials: jRCT1032230384.

### Clinical Protocols

https://jrct.mhlw.go.jp

### Author Declarations

Institutional Review Board of Toho University Ohashi Hospital Ethics Board H23024 H23 May 22, 2023

